# Acute kidney injury outcomes of COVID-19 confirmed patients in a Philippine tertiary hospital: A retrospective study

**DOI:** 10.1101/2024.02.05.24302369

**Authors:** Joberly Sayson, Reginald Kalaw, Bernadette Ecunar Ordona, Marissa Elizabeth Lim

## Abstract

The novel coronavirus (COVID-19) is known to be the fifth pandemic causing massive deaths worldwide. This virus has not only been deeply associated with acute respiratory distress, but also acute kidney injury (AKI). This study describes the baseline characteristics and various outcomes of AKI based on the KDIGO 2012 Clinical Practice Guidelines in patients hospitalized with COVID-19 at a Philippine tertiary hospital. A total of 195 patient records were retrospectively reviewed for the study. Of the 195 patients, 81(42%) patients developed AKI. Significant baseline characteristics included older age (56.28 + 14.12), presence of hypertension (p=0.004), diabetes mellitus (p=0.002), and cardiovascular disease (p=0.003). Also, the use of diuretics, inotropes and antibiotics were more prevalent in patients who developed AKI. Most of the patients who had AKI were categorized as stage 1 (49.38%). Mechanical ventilation was significantly (p<0.001) more prevalent in patients with AKI (20.99%) compared to patients without AKI (5.26%). There was significantly higher rates (p<0.001) of renal replacement therapy in patients with AKI (30.86%). Lastly, higher mortality rates were observed in patients with AKI (50.62%) versus patients without AKI (12.28%). Our study demonstrated that patients with COVID-19 can develop AKI and tend to have a poorer prognosis.

## Introduction

At the end of 2019, cases of a novel human pneumonia were first reported in Wuhan, China where patients reported symptoms of malaise, dry cough, dyspnea, and fever, with some requiring intensive care for severe and acute respiratory distress. The coronavirus disease of 2019 (COVID-19) quickly spread worldwide becoming the fifth recognized pandemic following the 1918 flu contagion [1]. The World Health Organization (WHO) reports that the number of COVID-19 cases has peaked at 767 million, with approximately 7 million deaths around the world since the start of the pandemic [2]. In the Philippines, the cumulative cases rose to more than 4 million with over 66,000 total deaths [2].

The coronaviruses are large, enveloped, single-stranded RNA viruses usually found in mammals such as dogs, cats, and humans. They cause neurologic, respiratory, and gastrointestinal diseases [3]. The Severe Acute Respiratory Syndrome Coronavirus-2 (SARS-CoV-2), in particular, is known to target the epithelial cells and pneumocytes of the nasal and bronchial areas through the viral structural spike (S) protein that binds to angiotensin-converting enzyme 2 (ACE 2) receptor [4]. Viral uptake is facilitated by cleaving the ACE 2 and activating the S protein of the virus [4]. Once within the cell, it promotes a cascade of events that trigger the inflammatory response, resulting in the disruption of the epithelial– endothelial integrity of the lungs. This eventually leads to acute respiratory distress syndrome (ARDS) [5].

In the kidneys, the virus can directly damage the renal tubular epithelium and podocytes through the ACE 2-dependent pathway. Viral invasion results in mitochondrial dysfunction, collapsing glomerulopathy, acute tubular necrosis, and protein leakage in the Bowman’s Capsule leading to acute kidney injury (AKI) [6].

These processes along with other factors, such as sepsis, hypovolemia, and cardiorenal syndrome, amplify the insult to the kidneys [7]. COVID-19 is also known to activate cytokine release syndrome (CRS), a phenomenon associated with the release of pro-inflammatory cytokines, such as interleukin-6, which could lead to systemic endothelial injury further impacting the kidneys [8]. Another mechanism that is known to be contributory is the organ crosstalk occurring between the cardiopulmonary system and the kidneys [9]. Hypoxemia resulting from ARDS and hypoperfusion due to COVID–19–related cardiac pathology may result in further damage to the renal tubules [10].

A meta-analysis of 44 studies conducted by Sabaghian et al reported AKI incidence to be as high as 56.2% in hospitalized COVID-19 patients and 75.5% after admission. The median age was reported to be 63 years, and over 40% had an existing comorbidity, such as CKD, chronic obstructive pulmonary disease, hypertension, diabetes, and tumors [11]. Proteinuria of any degree, hematuria of any degree, elevated baseline blood urea nitrogen (BUN), serum creatinine, peak serum creatinine > 133 μmol/l, and AKI over stage 2 were associated with in-hospital death after adjusting for age, sex, disease severity, comorbidities, and lymphocyte count [11]. Various studies have established the intrinsic association of AKI with poor prognosis (i.e., up to 83% at 5 years) and higher risk for mortality immaterial of pre-AKI baseline (hazard ratio [HR] 1.08 to 4.59) or recovery of renal function (HR 1.08 to 5.75) [12]. However, in patients with COVID-19 and AKI, there appears to be a 13- fold increased risk of mortality [13] with an estimated overall hospital mortality rate of 66.2% [11]. Among the several reasons for higher mortality were severe sepsis-related multi-organ failure, direct kidney involvement, and acute respiratory distress syndrome [14].

AKI in some COVID–19–confirmed patients may prompt the need for renal replacement therapy (RRT) or even a kidney transplant. The impact of these outcomes is more apparent in resource-limited settings, such as in the Philippine healthcare system [15]. AKI in COVID-19 encumbers the healthcare system by directly affecting the efficient utilization of hemodialysis machines, staff, and other hospital resources. Also, long-term monitoring is needed to avoid the progression of AKI to chronic kidney disease (CKD) and reduce the associated mortality in high-risk populations [15]. As such, it is important to recognize means to promptly identify and prevent further renal injury and facilitate recovery of renal function in patients with and who are recovering from COVID-19 [16].

In this study, we aimed to determine the clinical outcomes of patients who developed AKI in a tertiary hospital setting in the Philippines based on the Kidney Disease Improving Global Outcomes (KDIGO) 2012 Clinical Practice Guidelines AKI classification system. We sought to identify the baseline demographic profile of COVID-19 patients and determine the outcome in those who develop AKI and at what stage could lead to poorer outcomes. With this, we hope that our clinicians be guided in their clinical approach and prognostication of patients with greater risk for AKI to reduce or shorten the need for RRT and avoid complications like prolonged hospitalizations, development of CKD, as well as death.

## Materials and methods

### Study design

This is a retrospective cohort study that included a review of charts of all confirmed COVID-19 patients based on the result of their reverse transcription polymerase chain reaction (RT-PCR) test from 1 March 2020 to 31 December 2020. Baseline data, including age, co-morbidities, admission serum BUN and creatinine, urinalysis, and urine output monitoring were obtained. Data on medications during the hospital stay of the patients were also gathered. Outcomes such as discharge, death, and the need for assisted ventilation (e.g., high-flow nasal cannula, or mechanical ventilation) were collected **(Fig. 1)**.

**Figure 1.**
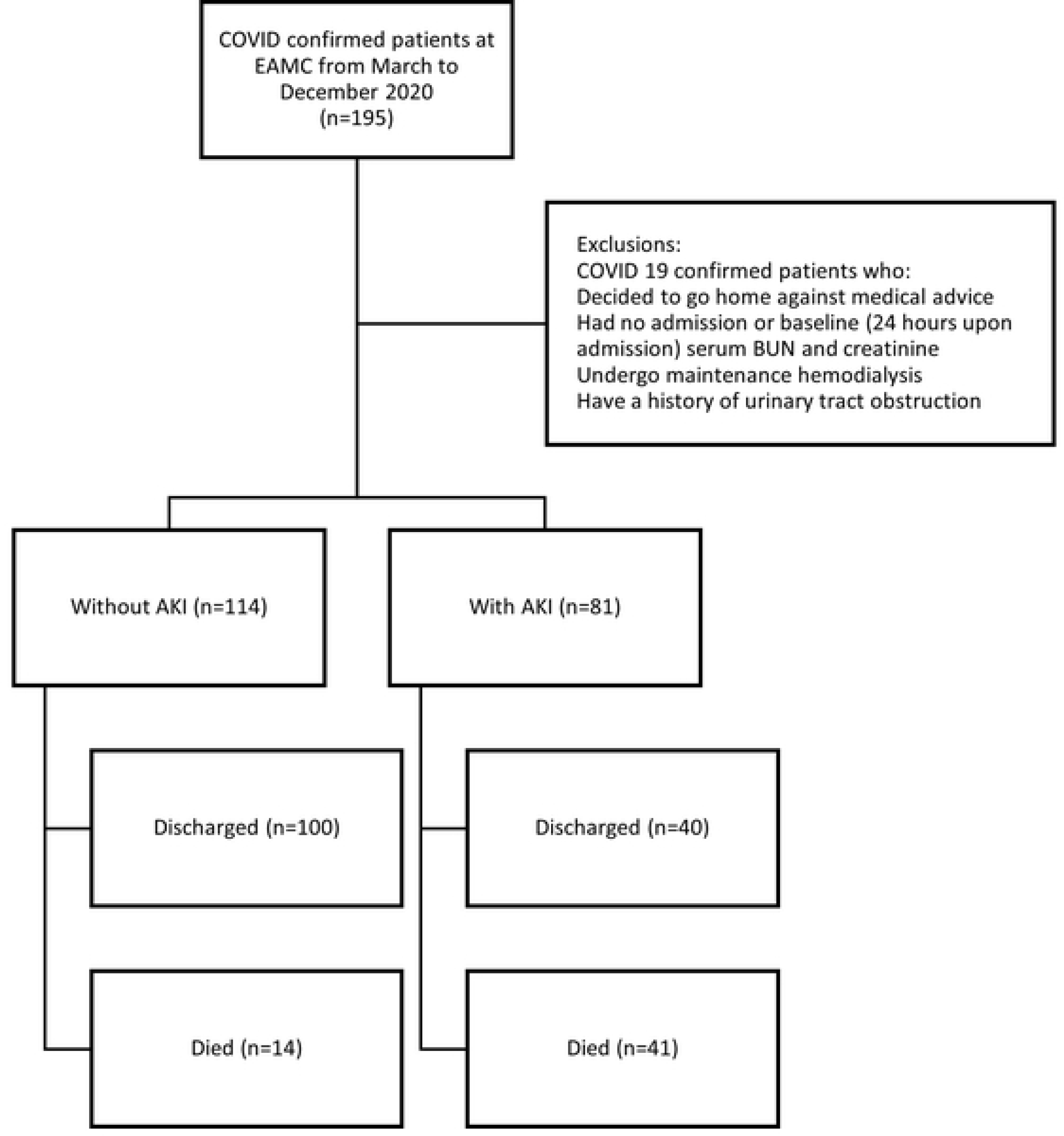
Subject selection process This figure outlines a summary of our study’s inclusion and exclusion process and demonstrates the clinical outcomes of COVID-19 patients admitted at the EAMC.

Patients who decided to go home against medical advice, had no baseline (within 24 hours upon admission) serum BUN and creatinine measurements, had undergone maintenance hemodialysis before admission, and had a history of urinary tract obstruction before admission were excluded.

### Sampling and sample size

A minimum of 185 patients were required for this study based on a 3.1% prevalence of AKI among patients with COVID-19 disease [17], and a 5% level of significance, and a 2.5% desired width of a confidence interval were set. A purposive sampling technique was employed with a total of 195 patients enrolled.

### Data gathering and ethical considerations

The study was submitted to the East Avenue Medical Center (EAMC) Institutional Ethics Review Board (IERB) for approval prior to the actual data collection and was conducted in compliance with the Philippine Data Privacy Act of 2012. A waiver of informed consent was requested and granted by the IERB. Only the primary investigator was involved in the collection of data. Patients included in the study were anonymized in the data collection form and only the case number and code numbers were used. General data, clinical features and laboratory results were extracted from the charts and recorded. The data gathered were then stored in a password-protected computer and stored in a lock and key cabinet. Data will be disposed via the secure delete feature as included in the operating system of the computer and documents used will be subjected to paper shredding 3 years after study initiation.

The list of COVID–19–confirmed patients from March 2020 to December 2020 was obtained from the Public Health Unit of a Philippine tertiary hospital, the EAMC in Quezon City. After, the charts of these patients were retrieved from the Medical Records for data collection. Baseline demographics such as age, gender, and co-morbidities such as hypertension, diabetes mellitus, and cardiac diseases, were documented. Serum BUN and creatinine levels determined within 24 hours of admission, as well as AKI findings during their hospital stay, were obtained, and classified based on the criteria of the KDIGO guidelines. Urinalysis and urine output monitoring were part of the data collection form. Medications given to the patients while admitted were likewise noted.

Patients were classified according to whether they developed AKI, whether they were discharged, expired, whether they required a high-flow nasal cannula or a mechanical ventilator, and their length of stay at the hospital recorded.

### Data processing and statistical analysis

Descriptive statistics were used to summarize the demographic and clinical characteristics of the patients. Frequency and proportion were used for categorical variables, median and interquartile range for non-normally distributed continuous variables and mean and SD for normally distributed continuous variables. An independent sample T-test, Mann-Whitney U test, and Fisher’s Exact/Chi-square test were used to determine the difference in mean, rank, and frequency, respectively, between the control and experimental groups. All statistical tests were two-tailed tests. The Shapiro-Wilk test was used to evaluate the normality of the continuous variables. Missing values were neither replaced nor estimated. The null hypotheses were rejected at a 0.05 level of significance. Lastly, the Stata 13.1 program was used for data analysis.

## Results

Patients with AKI were significantly older (56.28 ± 14.12; P<0.001) compared with patients without AKI (47.91 ± 14.72). There was no significant difference in the rates of AKI between male and female genders. Additionally, comorbidities, such as hypertension (p=0.004), diabetes mellitus (p=0.002), and cardiovascular disease (p=0.003), were significantly more prevalent in patients with AKI compared to patients without AKI, at 58.02% vs. 36.84%, 32.1 vs. 13.16%, 20.99 vs. 6.14%, respectively. Baseline creatinine clearance based on the Chronic Kidney Disease Epidemiology Collaboration (CKD-EPI) criteria showed that an estimated glomerular filtration rate (eGFR) of >90 (p<0.001) was significantly associated with AKI. Diuretics (p=0.001), inotropes (p=0.001), and the use of antibiotics were more prevalent in patients who developed AKI vs. non-AKI patients (34.57% vs. 13.16%, 20.99% vs. 5.26% and 100% vs. 85.09%, respectively).

Table 2 shows urinalysis results in patients with AKI vs. non-AKI, namely, the macroscopic and microscopic findings, chemical tests, and presence of crystals. Microscopic analysis reveals that AKI patients had significantly higher rates of hazy to turbid urine (p=0.042) vs. non-AKI patients. The presence of blood was likewise significantly higher (p=0.047) in those with AKI vs. non-AKI. The existence of bacteria (p=0.038) and amorphous urates (p=0.001) between AKI and non-AKI patients were also significantly different.

**Table 1.**
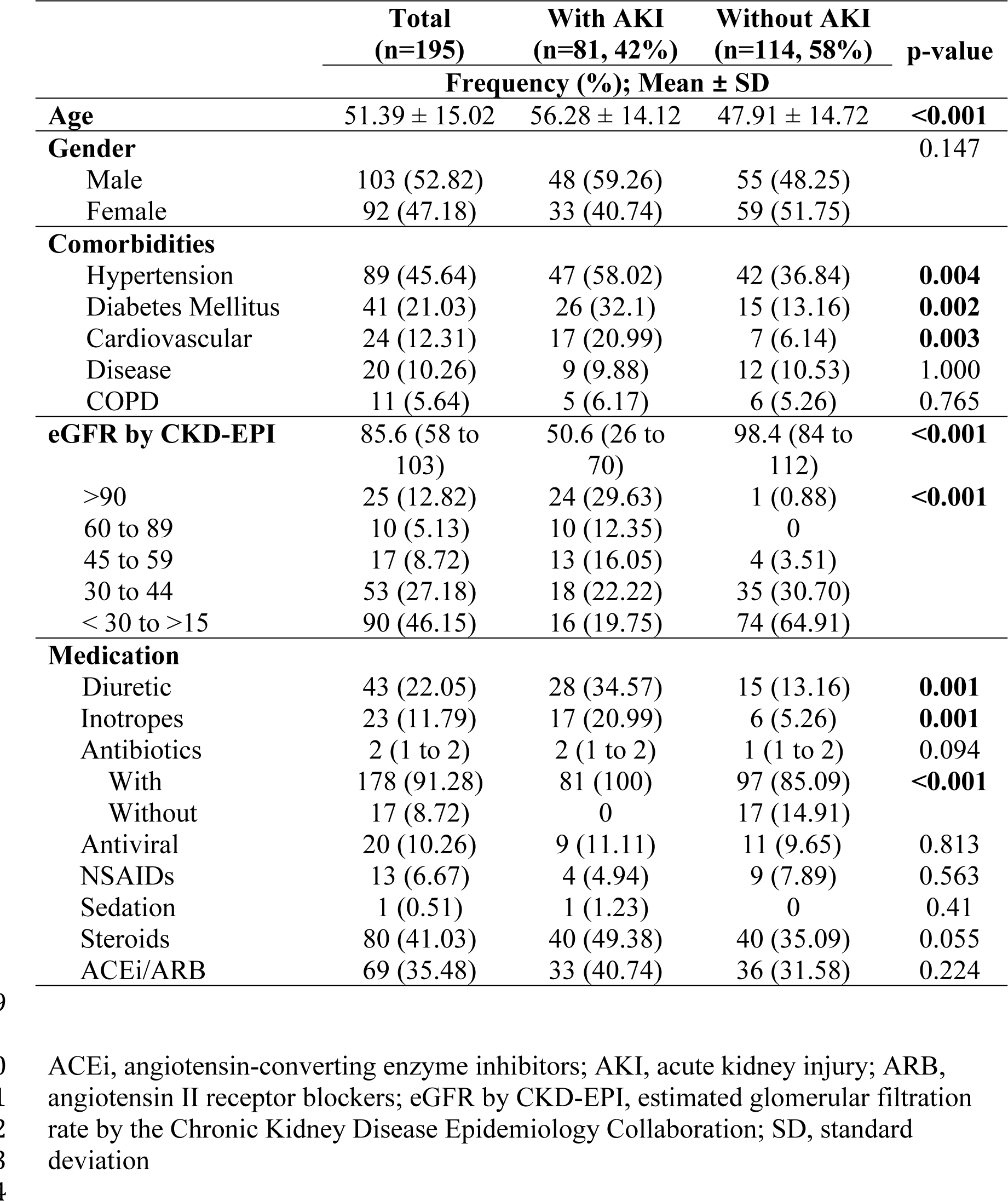
Baseline Characteristics of COVID-19 Confirmed Patients.

**Table 2.**
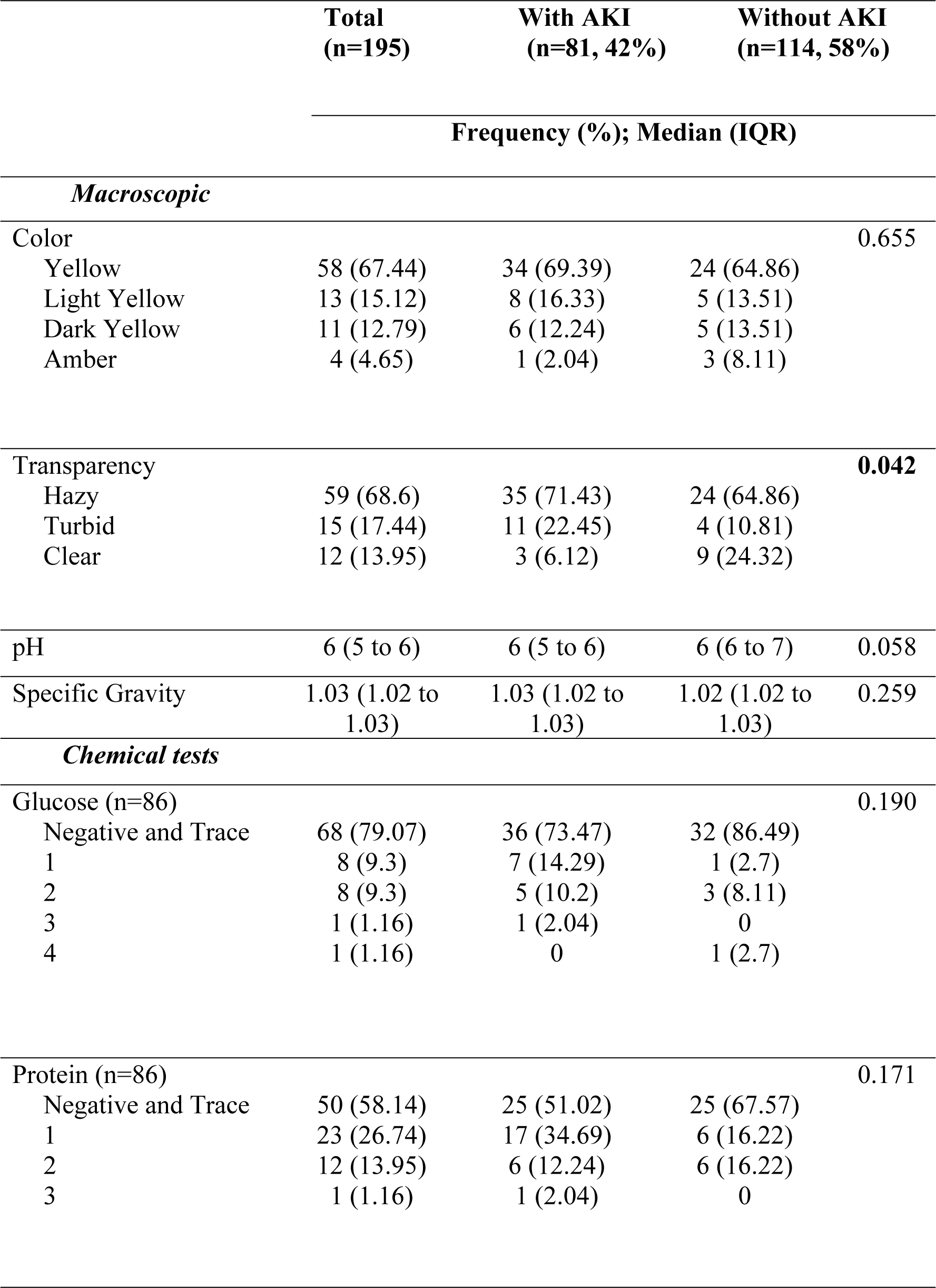
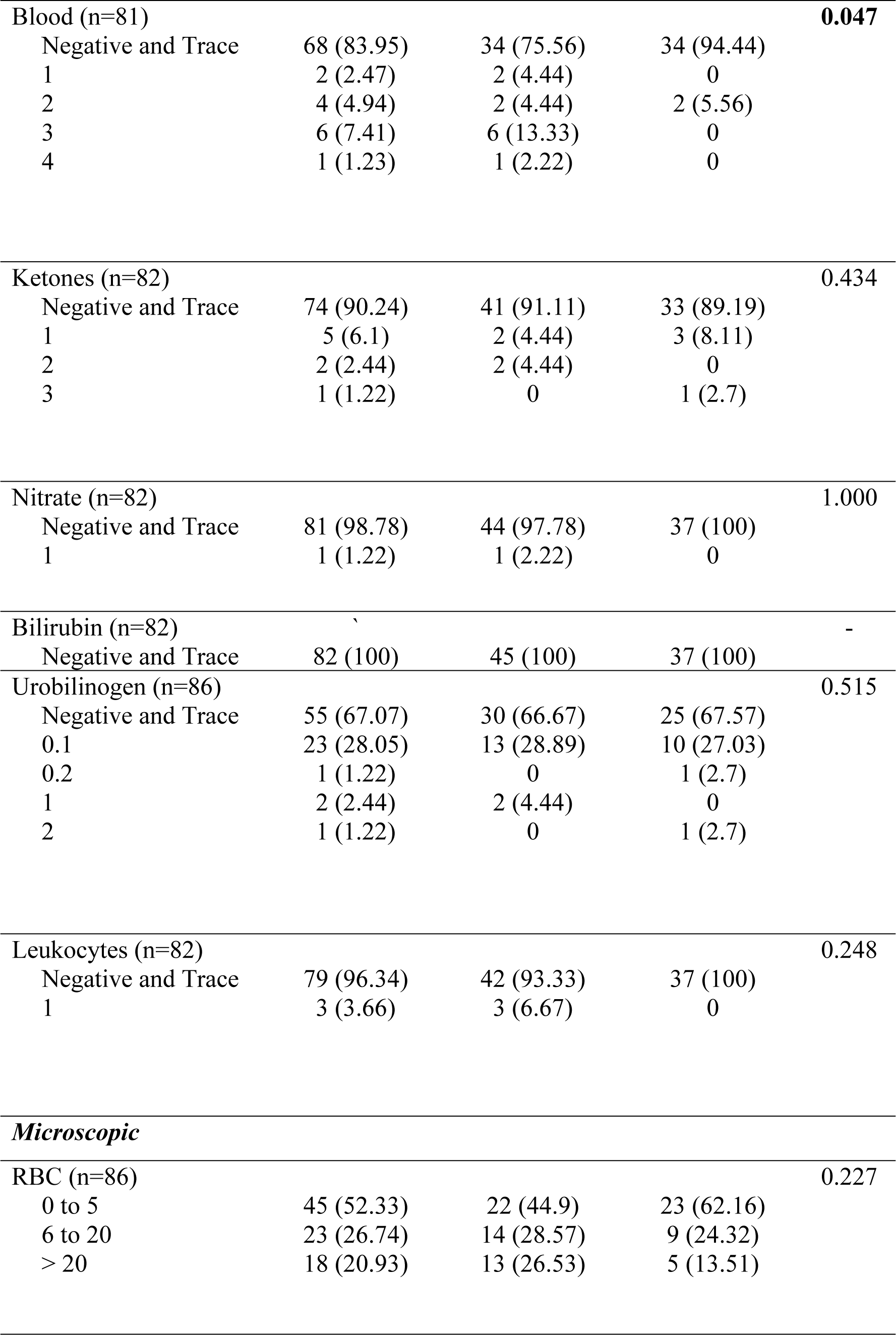
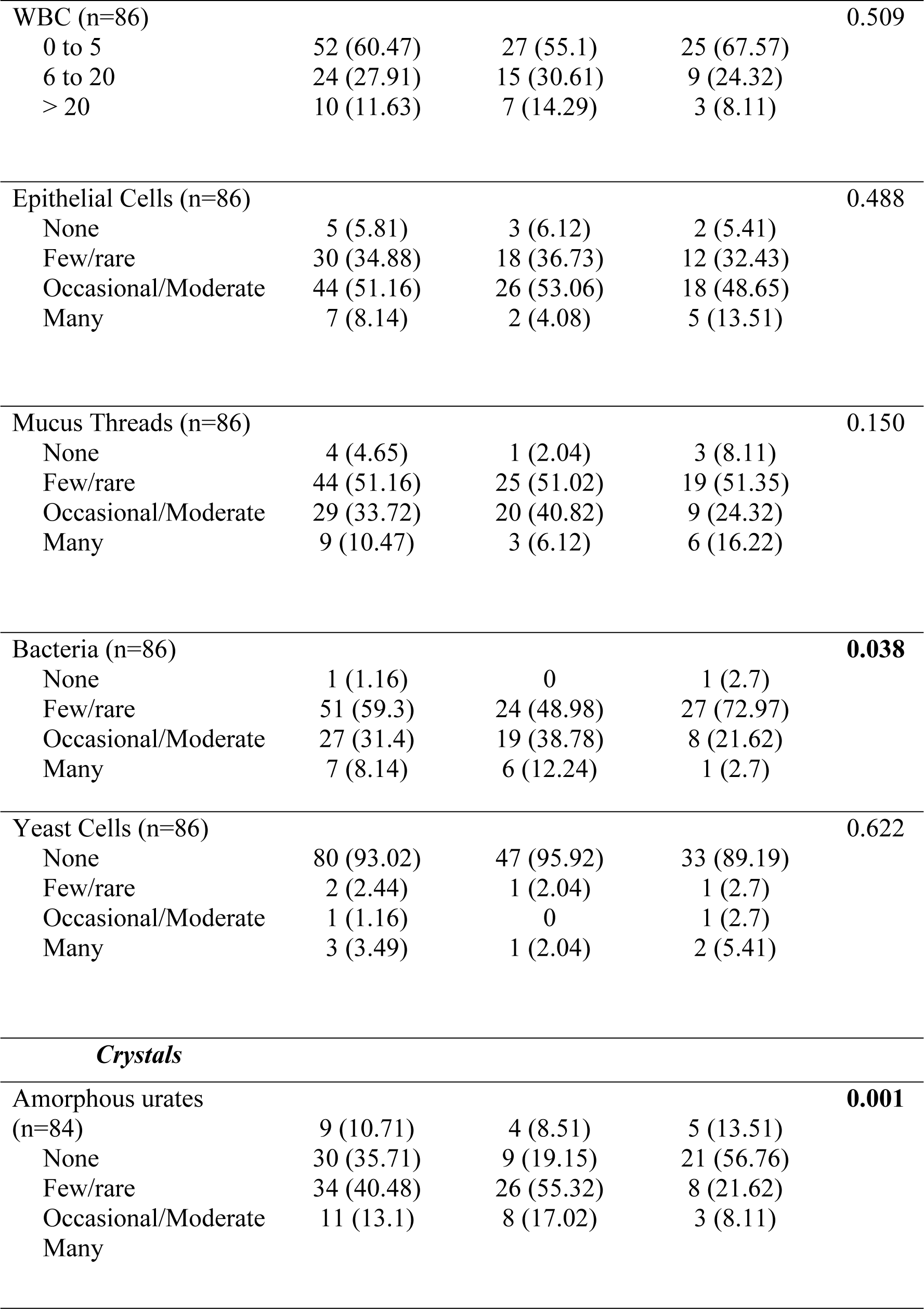
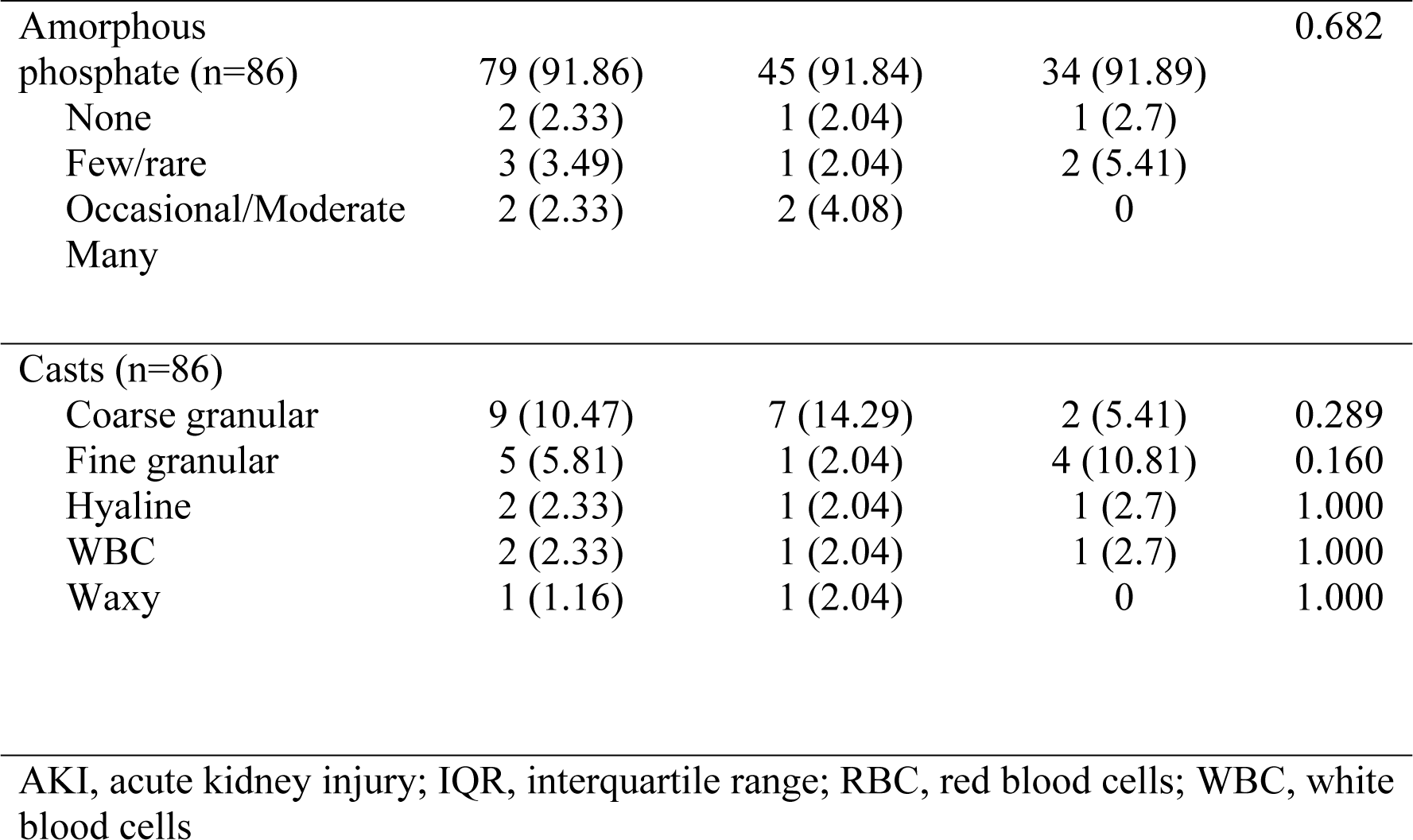
Urinalysis Results of COVID-19 Confirmed Patients.

Table 3 outlines that out of the 195 subjects reviewed, 81 developed AKI, the majority of whom developed stage 1 AKI. Table 4 shows the different outcomes in patients who developed AKI vs. those who did not develop AKI. There was significantly (p<0.001) greater mortality in patients with AKI (50.62%) vs. patients without AKI (12.28%). The length of hospital stay was not significant between AKI and non-AKI patients with an average hospital stay of 21 days. There was no significant difference among patients hooked to nasal cannula, facemask, or high-flow nasal cannula between AKI and non-AKI individuals. Mechanical ventilation was significantly (p<0.001) more prevalent in patients with AKI (20.99%) compared to patients without AKI (5.26%). There were significantly (p<0.001) more RRT cases in patients with AKI (30.86%) vs. patients without AKI (1.75%). The two patients who had undergone RRT without the development of AKI were those who had undergone isolated ultrafiltration while on hemoperfusion.

**Table 3.**
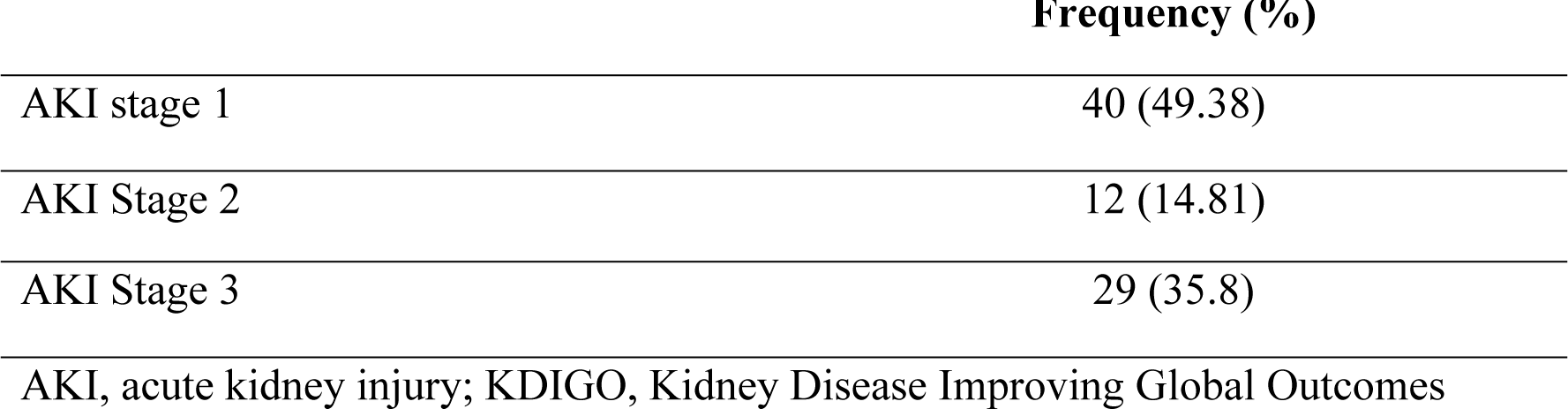
Frequency of Renal Function According to Staging of AKI Based on KDIGO 2012 (n=81)

|  | Frequency (%) |
| --- | --- |
| AKI stage 1 | 40 (49.38) |
| AKI Stage 2 | 12 (14.81) |
| AKI Stage 3 | 29 (35.8) |

AKI, acute kidney injury; KDIGO, Kidney Disease Improving Global Outcomes

**Table 4.** Assessment of Outcome of Patients With AKI vs. Those Without.

|  | **Total**<br>**(n=195)** | **With AKI**<br>**(n=81, 42%)** | **Without AKI**<br>**(n=114,**<br>**58%)** | **p-value** |
| --- | --- | --- | --- | --- |
|  | **Frequency (%); Median (IQR)** |  |  |  |
| Mortality |  |  |  | **<0.001** |
| Expired | 55 (28.21) | 41 (50.62) | 14 (12.28) |  |
| Discharged | 140 (71.79) | 40 (49.38) | 100 (87.72) |  |
| Length of Hospital Stay | 21 (12 to 27) | 20 (8 to 26) | 21 (15 to 28) | 0.053 |
| Morbidity |  |  |  |  |
| Nasal cannula | 74 (47.95) | 31 (38.27) | 43 (37.72) | 1.000 |
| Facemask | 28 (14.36) | 15 (18.52) | 13 (11.4) | 0.214 |
| High flow nasal cannula | 10 (5.13) | 5 (6.17) | 5 (4.39) | 0.744 |
| Mechanical ventilation | 37 (18.97) | 26 (32.1) | 11 (9.65) | **<0.001** |
| Renal replacement therapy |  |  |  | **<0.001** |
| Yes | 27 (13.85) | 25 (30.86) | 2 (1.75) |  |
| No | 168 (86.15) | 56 (69.14) | 112 (98.25) |  |

AKI, acute kidney injury; IQR, interquartile range

Table 5 demonstrates that there were more mortalities in stage 3 AKI while more discharges were observed in stage 1 AKI. The length of hospital stay was similar for the three stages. Most patients who had undergone RRT are in AKI stage 3, the seven patients who underwent RRT for stage 1 and stage 2 were patients who had undergone hemoperfusion, overlapping with hemodialysis.

**Table 5.** Assessment of Outcomes in Patients Based on AKI Stage.

|  | **AKI Stage 1** | **AKI Stage 2** | **AKI Stage 3** |
| --- | --- | --- | --- |
|  | **(n=40)** | **(n=12)** | **(n=29)** |
|  | **Frequency (%); Median (IQR)** |  |  |
| Mortality |  |  |  |
| Expired | 15 (37.5) | 5 (41.67) | 21 (72.41) |
| Discharged | 25 (62.5) | 7 (58.33) | 8 (27.59) |
| Length of Hospital Stay | 21 (8 to 26) | 24 (14 to 30) | 10 (4 to 25) |
| Renal replacement therapy |  |  |  |
| Yes | 5 (12.5) | 2 (16.67) | 18 (62.07) |
| No | 35 (87.5) | 10 (83.33) | 11 (37.93) |

AKI, acute kidney injury; IQR, interquartile range

## Discussion

Our study has outlined the number of patients who developed AKI, their baseline characteristics, outcomes, and those who underwent RRT in patients admitted for COVID-19 at the EAMC from March to December 2020. The EAMC is one of the largest hospitals to admit COVID-19 patients in Metro Manila. Of the 195 patients included in the study, 81 (41.5%) of them developed AKI. These findings mirror the results of a US study where 49.3% of patients who had COVID-19 developed AKI [18]. Out of the 81 patients in our study who developed AKI, a majority (49.38%) developed stage 1, followed by the development of stage 3 which is 35.8%, and lastly by stage 2 which is 14.81%.

Several studies have confirmed that age is a factor for the development of AKI in COVID-19. For instance, in the study of Nimkar and colleagues, it was shown that the median age of the patients was 71 years [19] while it was 61 years old for the study of Fominsky et al [20]. Most individuals admitted for COVID-19 at our center tended to be older (56.28 ± 14.12). The significant comorbidities include hypertension, diabetes mellitus and cardiovascular diseases (58.02%, 32.1% and 20.99% respectively). This is consistent with the study conducted by Sargiacomo et al associating worse outcomes with these comorbid illnesses and they represent risk factors for the development of AKI [21].

Several factors may predispose the critically ill to the development of AKI in COVID-19. Hypoperfusion of the kidneys secondary to shock resulting from sepsis, hypovolemia, or cardiogenic etiologies, especially in patients with underlying cardiac disease, may be one of the causes [22]. On the other hand, pre-existing COPD was not seen as a predisposing factor in our study. As for medications used at the time of admission, diuretics, inotropes, and antibiotics tended to be more significantly prevalent in patients who developed AKI compared with other medications such as steroids, ACEi/ARBs, antivirals and NSAIDs. In a study by Nadim et al, the medications that were found to be contributory to AKI development include antibiotics, contrast media, and NSAIDs. They attributed the development of AKI acute tubular injury [23].

Cheng et al observed that hematuria and proteinuria were associated with a threefold and almost twofold increase in the risk of AKI in COVID-19 patients; however, this was not replicated in our study [24]. We do note that urine transparency and the microscopic presence of red blood cells (RBCs) are associated with AKI. In a Turkish study, histopathological findings in kidneys of COVID-19 patients include the presence of viral particles in the proximal tubules, podocytes with varying degrees of acute tubular necrosis and loss of brush borders, vacuolar degeneration and the accumulation of RBCs [25], possibly supporting these findings.

Patients who developed AKI tended to have poorer prognosis with most of them succumbing to death (50.62% vs. 12.28%). This is more apparent in those who developed AKI stage 3 (72.41% vs. 27.59%). Like the systematic review by Ali et al, mortality was significantly higher in patients with AKI [26]. The length of hospital stay appears to have no bearing on the development of AKI in COVID-19.

Organ crosstalk appears to be the mechanism implicated in the involvement between alveolar and tubular damage as they are both contributory to interleukin-6 (IL-6) overproduction [9]. This may be the underlying reason for the association between mechanical ventilation and AKI. Aside from organ crosstalk, pulmonary inflammation may lead to the release of pro-apoptotic and pro-inflammatory substances that may lead to AKI [22].

Most importantly, out of our 81 patients who developed AKI, 25 underwent RRT (30.9%) where 18 had stage 3 AKI while seven underwent RRT while undergoing hemoperfusion. Because of this, we reiterate the importance of close monitoring in patients with AKI to prevent the need for RRT.

We acknowledge that our study has some limitations. Firstly, our analysis was limited to a retrospective cohort and as such, comes with the limitations inevitably linked with this type of study. Secondly, laboratory findings, such as urinalysis results were not uniformly available in all patients, while serum creatinine and blood urea nitrogen (BUN) levels were not regularly monitored. Lastly, no post discharge follow-up was conducted in our study.

## Conclusions

Our study demonstrated that patients with COVID-19 can develop AKI and tend to have a poorer prognosis, including death, especially at stage 3 AKI. Older age, the presence of hypertension, diabetes mellitus, and cardiovascular disease can be risk factors for the development of AKI as well as the use of diuretics, inotropes, and antibiotics. The development of AKI in COVID-19 patients can lead to RRT and monitoring renal function after recovery from AKI is important to avoid progression to chronic renal failure.

## Data Availability

East Avenue Medical Center Medical Records

## Acknowledgments

We thank our mentors and colleagues at the Section of Nephrology of the EAMC for their support and guidance.

